# Interim evaluation of Google AI forecasting for COVID-19 compared with statistical forecasting by human intelligence in the first week

**DOI:** 10.1101/2020.12.16.20248358

**Authors:** Junko Kurita, Tamie Sugawara, Yasushi Ohkusa

## Abstract

**Background:** Since June, Google (Alphabet Inc.) has provided forecasting for COVID-19 outbreak by artificial intelligence (AI) in the USA. In Japan, they provided similar services from November, 2020.

**Object:** We compared Google AI forecasting with a statistical model by human intelligence.

**Method:** We regressed the number of patients whose onset date was day *t* on the number of patients whose past onset date was 14 days prior, with information about traditional surveillance data for common pediatric infectious diseases including influenza, and prescription surveillance 7 days prior. We predicted the number of onset patients for 7 days, prospectively. Finally, we compared the result with Googles AI-produced forecast. We used the discrepancy rate to evaluate the precision of prediction: the sum of absolute differences between data and prediction divided by the aggregate of data.

**Results:** We found Google prediction significantly negative correlated with the actual observed data, but our model slightly correlated but not significant. Moreover, discrepancy rate of Google prediction was 27.7% for the first week. The discrepancy rate of our model was only 3.47%.

**Discussion and Conclusion:** Results show Googles prediction has negatively correlated and greater difference with the data than our results. Nevertheless, it is noteworthy that this result is tentative: the epidemic curve showing newly onset patients was not fixed.

## Introduction

In Japan, the COVID-19 outbreak had two peaks until October. Data show that it worsened again in November, 2020 (Figure 1) provided by Ministry of Health, Labour and Welfare (MHLW) [1]. Even though the first peak around April is explainable by voluntary restriction of going out (VRG) [2], the second peak around July cannot be explained by any mathematical model to date.

**Figure 1:**
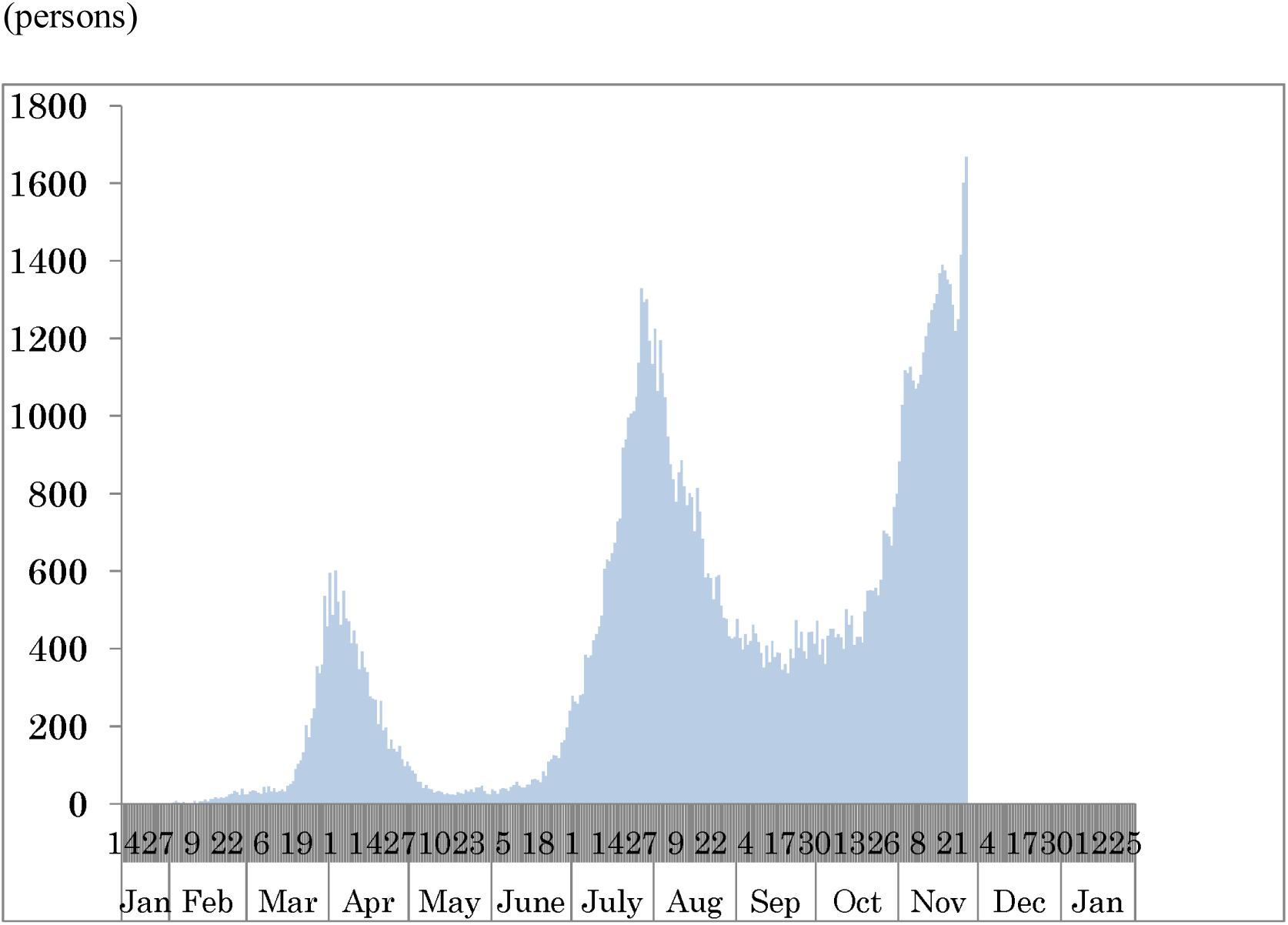
Epidemic curve of COVID-19 outbreak from January 14 to the end of November, 2020. Note: The line represents epidemic curve, which means the number of the newly onset patients each day. Data were obtained from MHLW, Japan.

Google (Alphabet Inc.) has provided forecasting by AI for the COVID-19 outbreak since May [3]. In Japan, Google started to provide similar services since November, 2020 [4]. Nevertheless, the manner by which and the quality with which the AI solves this problem of prediction which cannot be solved by human intelligence has not been evaluated to date. To elucidate those points, we undertook comparison of Google AI forecasting results to those obtained using a statistical model without AI.

Especially, we used all information about infectious diseases including National Official Sentinel Surveillance for Infectious Diseases (NOSSID), except for COVID-19 [5] and prescription surveillance (PS) [6–9], which were available for more than ten years before the COVID-19 outbreak occurred.

By MHLW in Japan, the incidence of infectious disease has been monitored at medical institutions through official national surveillance of infectious diseases based on the Law for the Prevention of Infectious Diseases and Medical Care for Patients of Infections. By NOSSID, cases of almost all common pediatric infectious diseases, except for influenza, have been reported from 3,000 sentinel medical institutions, collectively accounting for only about one-tenth of all pediatric medical institutions. For influenza, 2,000 internal medicine facilities were added as sentinels for monitoring. Moreover, because the number of patients with each infectious disease per sentinel in the prior week is announced once each week (every Friday at noon) at a nationwide level, this report has been published with a 7–10 day delay. The sentinel medical institutions for these diseases have specifically established pediatric hospitals and clinics. Therefore, these reports might not include information for adults and elderly patients; alternatively, they might heavily underreport that information.

Since 2007, PS has been developed through cooperation among research groups headed by Dr. Ohkusa and EM Systems Co. It has been operated by the Japan Medical Association, Japan Pharmaceutical Association, School of Pharmacy, Nihon University, and EM Systems Co. Ltd. It reports the estimated number of patients based on information of prescriptions filled at external pharmacies. In November 2020, approximately 13,000 pharmacies participated, collectively accounting for about 20% of all pharmacies in Japan.

This system mainly monitors prescriptions from hospitals or clinics that use Application Service Provider for medical claims in pharmacies through a safe internet connection. The collected data represent the number of prescriptions given, classified into therapeutic categories as described below. Therefore, the collected data include no personal information. Data related to the number of prescriptions are extracted automatically and are analyzed daily.

In this system, the numbers of patients were estimated from the numbers of prescriptions for neuraminidase inhibitors, anti-varicella-herpes-zoster virus (VZV) drugs, antibiotic drugs, antipyretic analgesics, and multi-ingredient cold medications by prefecture each day. The numbers of patients with neuraminidase inhibitors or anti-VZV drugs were classified by three age groups: children younger than 15 years old, adults younger than 64 years old, and elderly people over 65 years old and older. Moreover, antibiotics were classified into five types: penicillin, cephem, macrolide, new quinolone, and others [8]. These drugs were chosen to identify clusters of rash, fever, or digestive symptoms to detect bioterrorism attack, emerging/remerging diseases, and mass food poisoning. Particularly because anti-VZV drug is a drug for varicella and zoster, a cluster of this drug in adults with no cluster in children or elderly people represents a signal of small pox [6]. The following morning, these data are presented on a web page (http://prescription.orca.med.or.jp/syndromic/kanjyasuikei/).

## Methods

### Data

Information about the COIVD-19 outbreak was provided by MHLW [1]. The PS system estimates the numbers of patients by multiplying the reciprocal of the participation rate of pharmacies to PS. It also estimates the reciprocal of the proportion of external prescription in the prefecture to the total number of prescriptions in the prefecture. The numbers of patients who received neuraminidase inhibitors, anti-VZV drugs, antibiotic drugs, antipyretic analgesics, multi-ingredient cold medications, and antidiarrheal/ intestinal drugs at an external pharmacy are recorded.

We examined the short-term prediction of the respective incidences of influenza, RS virus infection (RS), pharyngoconjunctival fever (PCF), group A streptococcal pharyngitis (A-SP), varicella, hand, foot and mouth disease (HFMD), erythema infectiosum (EI), exanthem subitum (ES), herpangina, and mumps in NOSSID. Also, NOSSID provides the numbers of patients per sentinel per week as the incidence of each disease considered in this study, except for RS. For RS, NOSSID provides only the total number of patients per week. This information has been published officially every Friday at noon, reflecting the situation of the prior week. For the procedures explained below, we use the latest information of NOSSID from Saturday. Therefore, on Saturday and Sunday, we can use NOSSID information for the prior week or of earlier weeks.

### Estimation model

We first examined the association among the epidemic curve of the COVID-19 outbreak and the incidences from NOSSID and the information from PS. We regressed the number of patients whose onset date was day *t* on their lagged variables for two weeks, *t-1* to *t-k*, and the information of PS and NOSSID available one week before, on day *t-7*. The length of autoregressive parts, *k*, was determined to reach the highest the adjusted coefficient of determinant. As described above, NOSSID data are delayed by 10–14 days: the information available on day *t-7* were data for *t-17* through *t-21*. For estimation, we used data from May 1 to the end of October. About one month was necessary to fix the epidemic curve. Therefore, we did not use data for the last 30 days.

Google explained their procedure as AI deep learning for a SIR model, although details of the final model were not published [3]. They provided the number of cumulative confirmed cases, cumulative deaths, hospitalized patients, recovered and their 95% intervals. They have not shown a forecast of newly onset number of patients, meaning the epidemic curve. Among them, we specifically examined newly confirmed cases, which were defined as incremental cumulative confirmed cases at day *t*.

### Prediction

In our model, although the lagged variable of the number of newly onset patients for *t-1* to *t-k* can be predicted recursively, information from PS and NOSSID cannot be predicted. The maximum prediction period is therefore up to 7 days. We updated our prediction every day from November 21, 2020. However, for comparison with the Google prediction produced using the same information, we evaluated the first prediction on November 21 for one week. In other words, we did not use predictions using information related to November 22, 2020 or later. Therefore, we note that our prediction after November 22 was not the same as the prediction on November 21, even for the same day. By contrast, Google provided their prediction for 4 weeks from November 19, 2020.

### Evaluation of prediction

We evaluated the predictive capability of the two models by the discrepancy rate and correlation coefficient among predictions from the data. the weighted average of absolute value of difference among the actual data and forecasting divided by the actual data

Google published their prediction for four weeks from 21 November on 20 November. Prediction by our model started on 21 November. Therefore, it predicted for the period of 22–28 November. Predictions made during 22–28 November were of the period for evaluation.

As described earlier, the epidemic curve was expected to change over time within 30 days. However, we did not wait for such a longer time to evaluate predictions. Instead, we used the epidemic curve as of the end of November, which was three days after the last day of evaluation.

### Ethics

Data from PS were aggregated and de-linked from personal information related to patients, medical institutions, and pharmacies: these are anonymous data. Information from NOSSID and Google were published data. Therefore, no ethical issue was posed by the use of these data for this study.

## Results

The adjusted coefficients of determination indicate that the maximum length of the auto regression part, *k*, was 14. Figure 2 depicts the epidemic curve and fitted values based on estimation results of our statistical model within the sample. It is apparently almost perfectly fitted within the sample: the adjusted coefficient of determination was 0.9896.

**Figure 2:**
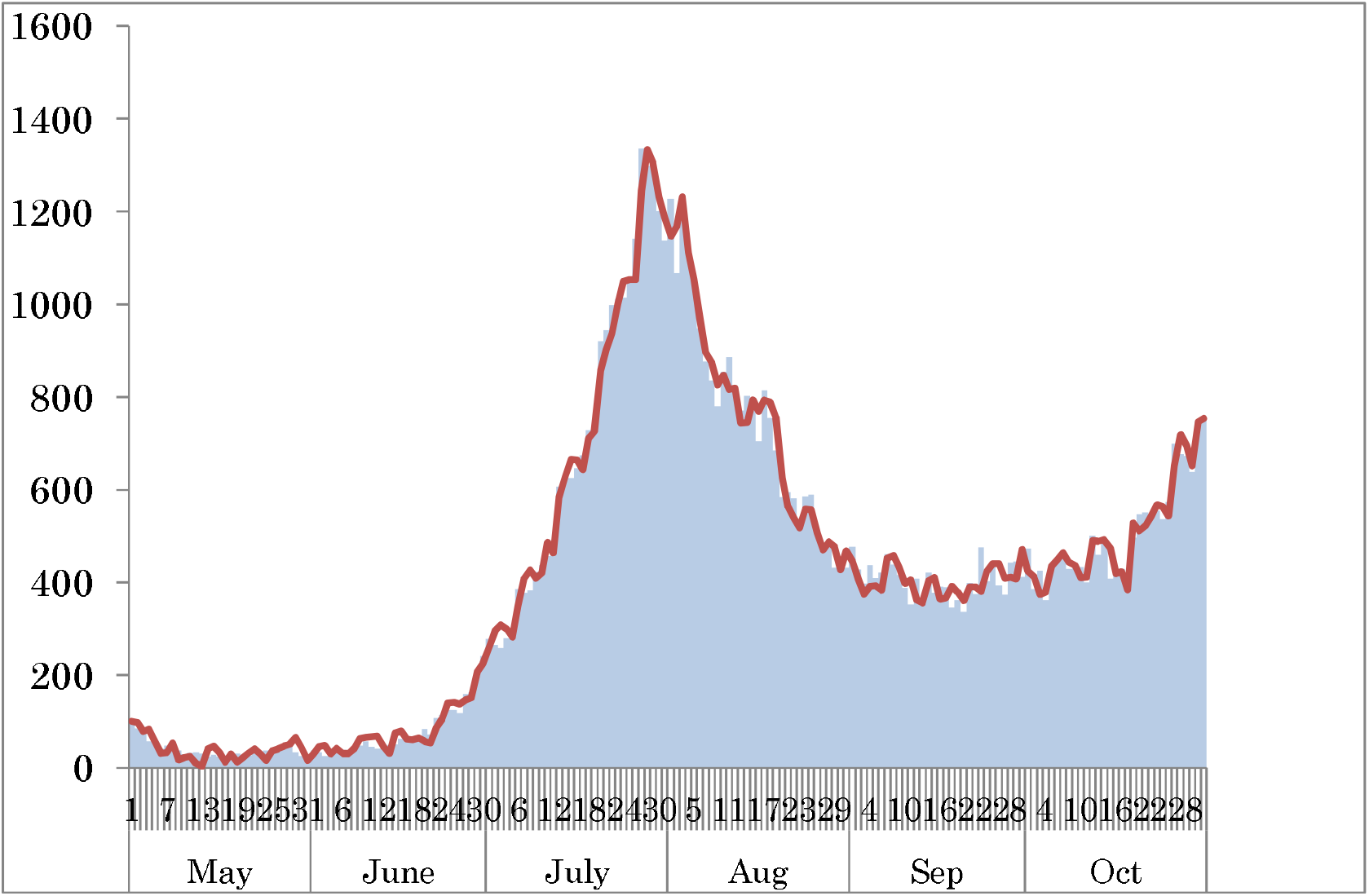
Epidemic curve and fitted values by the statistical model. Note: Data are from May 1 to the end of October. Bars represent the observed epidemic curve. The red line was fitted line by the statistical model. The estimated adjusted coefficient of determination was 0.9896.

Figure 3 depicts the observed epidemic curve as of the end of November and our prediction from November 20, 2020. It also showed the number of newly confirmed cases and Google’s prediction for that number. The discrepancy rate for the Google prediction was 27.7% for the first week: the discrepancy rate of our model was only 3.47%.

**Figure 3:**
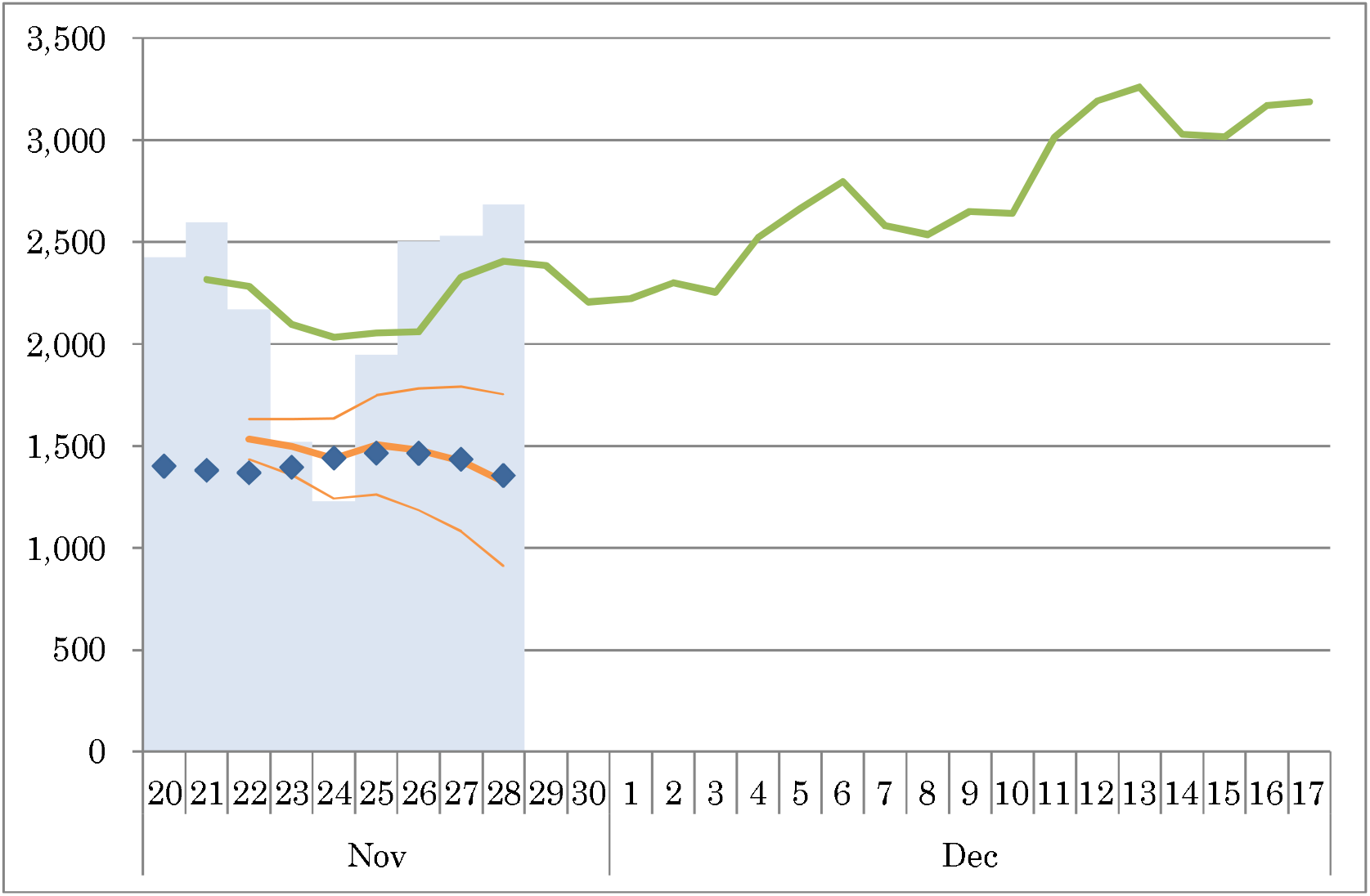
Epidemic curve, number of newly confirmed patients each day, and forecasting by Google AI and our statistical model. (patients) Note: Bars represent the numbers of newly confirmed patients; dots denote the numbers of newly onset patients as of the end of November. The green line represents Google’s AI prediction of the number of newly confirmed patients. The orange line shows predictions by our statistical model using information related to November 21. Thin lines were its 95%CI.

On the other hand,, correlation coefficients of the two predictions in the present study were -0.801-for Google and 0.3118 for ours. The *p*-values were 0.030 and 0. 0.496, respectively. Therefore, Google has significantly negative correlated, and our model show positive correlated but insignificant.

## Discussion

Results show that our model was superior to that of the Google AI prediction in terms of both of the discrepancy rate and correlation rate. As shown in Figure 3, the observed epidemic curve lay within the band of CI of our model’s prediction, no significant difference was found between the predicted and observed values. Conversely, CI of Google’s prediction for the newly confirmed cases was not provided, although it provided CI of prediction of the cumulative confirmed cases.

Although correlation coefficients are sometimes used for evaluation, they are insufficient to evaluate the gap separating data and prediction. It simply indicated whether two data were proportional or not. Therefore, it might be inappropriate for the evaluation of prediction because it is most important that the average of prediction was similar with the data. Therefore, we adopted the discrepancy rate for evaluation of prediction.

Because Google AI prediction depends on the mathematical model, it probably cannot explain several peaks of the COVID-19 outbreak because the mathematical model implies that the peak will be achieved by herd immunity when the proportion of the infected persons was higher than 1-1/*R*_0_. Actually, a study conducted in the US [3] used data in the upward phase before the first peak. In other words, they did not evaluate that the model can predict the peak. In the upward phase only, the mathematical model can explain the data well, but it does not have predictive power for the peak or reemergence of the outbreak. In this sense, their study appeared to be too early to evaluate the model.

Similarly, regarding forecasting in Japan, details were not disclosed in the manuscript, the Google AI probably used data after July, when the outbreak was downward sloping or almost stable. In this or a similar phase, any model can probably predict outcomes easily. In this sense, the Google AI forecasting model has not been evaluated yet for its prediction power for the peak. Conversely, our examined statistical model can explain the second peak around the end of July. Therefore, it might predict the third or later peaks.

As described above, the Google AI forecasting model has provided the number of newly confirmed cases rather than the number of onset patients, which means the epidemic curve. The series of the number of confirmed cases might be meaningless in terms of epidemiological perception because it might be affected by the testing capacity or strategy or the day of the week. For instance, the building capacity of testing might find a higher proportion of asymptomatic cases and might exaggerate the size of the outbreak. Actually, newly confirmed cases were fewer for the weekend and higher on the days after holidays or Sunday. It might result from the fact that fewer patients visit a doctor during the weekend or on holidays. However, the number of newly confirmed cases was always reported publicly and is very familiar to the general population. Moreover, the figure will not change over time.

Conversely, our model predicted the number of onset patients, meaning that it predicted the epidemic curve. It is therefore more appropriate from the perspective of epidemiology. However, because there were lags from onset to reporting and distribution of the report, the number of onset patients in the most recent days certainly fluctuates over time for two weeks or for one month. In other words, two weeks or one month duration is necessary to fix the epidemic curve. Therefore, precise prediction for the epidemic curve is extremely important. However, because it requires fixing for a long time, it was not published publicly and routinely and is not commonly available to the general public.

By the definition of AI, because the AI can use knowledge or a model used by human intelligence and because human intelligence cannot access the AI model in detail, AI can always provide better prediction than human intelligence. The problem remains of whether differences between predictions by an AI and human intelligence are useful or not.

In general, AI cannot predict new situation which was not included teaching data [11]. Because neural network adjust their parameter in the region of [0,1]. It cannot set some parameter as higher than one or lower than zero. In other words, AI was a kind of average in the teaching data though its procedure was complex. Therefore, AI cannot expect outside the set of teaching data. However, if the situation was included in the set f teaching data, it follows usual pattern, and thus prediction should be easily even by human being. Forecasting should be important in the newly emerging situation which was unusual and not included teaching data. In this case, AI may not predict better than human intelligence.

Although definitions of predicted variables for Google’s AI and our model were not the same, our predictive model was shown to be more precise than Google’s AI model. The present study has some limitations. First, although we evaluate two predictions in the first week, evaluations might be different for a longer time or a different phase of the outbreak. Therefore, we must continue to evaluate the two predictions over a longer time.

Secondly, the epidemic curve predicted by our model was not fixed until one month later. Therefore, evaluation for the prediction of our model might change up to one month. The obtained result should be regarded as tentative. By contrast, the evaluation of Google’s prediction will not change.

## Conclusion

We demonstrated that our model was more appropriate than Google’s for the first week. However, it is noteworthy that this result is tentative: not conclusive. We must monitor these two predictions’ respective performances carefully.

The present study is based on the authors’ opinions: it does not reflect any stance or policy of their professionally affiliated bodies.

## Data Availability

Japan Ministry of Health, Labour and Welfare. Press Releases.
Japan: COVID-19 Public Forecasts - Data Studio

https://www.mhlw.go.jp/stf/newpage_10723.html

https://datastudio.google.com/reporting/8224d512-a76e-4d38-91c1-935ba119eb8f&#12288;

## Acknowledgments

We acknowledge the great efforts of all staff at public health centers, medical institutions, and other facilities who are fighting the spread and destruction associated with COVID-19.

## Reference

1. Japan Ministry of Health, Labour and Welfare. Press Releases. https://www.mhlw.go.jp/stf/newpage_10723.html (in Japanese) [accessed on November 24, 2020]

2. Kurita J, Sugawara T, Ohkusa Y. Mobility data can explain the entire COVID?19 outbreak course in Japan. medRxiv 2020.04.26.20081315; doi: https://doi.org/10.1101/2020.04.26.20081315

3. Arik S, Li C, Yoon J, Sinha R, Epshteyn A, Le LT, Menon V, Singh S, Zhang L, Nikoltchev M, Sonthalia YK, Nakhost H, Kanal E, Pfister T. Interpretable Sequence Learning for COVID-19 Forecasting. NeurIPS 2020. https://research.google/pubs/pub49500/

4. Japan: COVID-19 Public Forecasts - Data Studio https://datastudio.google.com/reporting/8224d512-a76e-4d38-91c1-935ba119eb8f (in Japanese) [accessed on 21 November, 2020]

5. Hashimoto S, Murakami Y, Taniguchi K, et al. Annual incidence rate of infectious diseases estimated from sentinel surveillance data in Japan. J Epidemiol. 2003;13:136–41.

6. Sugawara T, Ohkusa Y, Ibuka Y, Kawanohara H, Taniguchi K, Okabe N. Real-time prescription surveillance and its application to monitoring seasonal influenza activity in Japan. J Med Internet Res 2012;14(1):e14.

7. Sugawara T, Ohkusa Y, Kawanohara H, Kamei M. Short Term Prediction of Infectious Diseases Patients from Prescription Surveillance. Journal of Biosciences and Medicines 2018. 6(9): 61–8.

8. Ohkusa Y, Sugawara T, Kawanohara H, Kamei M. Evaluation of the global action plan on antimicrobial resistance in Japan during its first eighteen months. Drug Discov Ther. 2018; 12(3):182–184.

9. Sugawara T, Ohkusa Y, Kawanohara H, Kamei M. Prescription surveillance for early detection system of emerging and reemerging infectious disease outbreaks. Biosci Trends. 2018;12:523–5.

10. Sugawara T, Ohkusa Y, Kawanohara H, Kamei M. Short Term Prediction of Infectious Diseases Patients from Prescription Surveillance. Journal of Biosciences and Medicines 2018. 6:61–8.

11. D’Amour A, et al. Underspecification presents challenges for credibility in modern machine learning. 2011.03395v2 [cs.LG]24Nov2020

